# Cardiopulmonary Exercise Testing in a Prospective Multicenter Cohort of Older Adults: The Study of Muscle, Mobility and Aging (SOMMA)

**DOI:** 10.1101/2023.09.25.23296120

**Authors:** Cody Wolf, Terri L. Blackwell, Eileen Johnson, Nancy W. Glynn, Barbara Nicklas, Stephen B. Kritchevsky, Elvis A. Carnero, Peggy M. Cawthon, Steven R. Cummings, Frederico G. S. Toledo, Anne B. Newman, Daniel E. Forman, Bret H. Goodpaster

**Affiliations:** Department of Health and Physical Activity, University of Pittsburgh, Pittsburgh, Pennsylvania, USA; San Francisco Coordinating Center, California Pacific Medical Center Research Institute, San Francisco, California, USA; Department of Epidemiology, University of Pittsburgh, Pittsburgh, Pennsylvania, USA; Department of Internal Medicine-Gerontology and Geriatric Medicine, Wake Forest University School of Medicine, Winston-Salem, North Carolina, USA; Translational Research Institute, AdventHealth, Orlando, FL, USA; Department of Epidemiology and Biostatistics, University of California, San Francisco, California, San Francisco, California, USA; Department of Medicine-Division of Endocrinology and Metabolism, University of Pittsburgh School of Medicine, Pittsburgh, Pennsylvania, USA; Department of Medicine (Geriatrics and Cardiology) University of Pittsburgh; Geriatrics Research, Education and Clinical Care (GRECC), VA Pittsburgh Healthcare System, Pittsburgh, PA

**Keywords:** cardiorespiratory fitness, aging, frailty, muscle, VO2peak

## Abstract

**BACKGROUND:** Cardiorespiratory fitness (CRF) measured by peak oxygen consumption (VO2peak) declines with aging and correlates with mortality and morbidity. Cardiopulmonary Exercise Testing (CPET) has long been the criterion method to assess CRF, but its feasibility, efficacy and reliability in older adults is unclear. The large, multicenter Study of Muscle, Mobility and Aging (SOMMA) employed CPET to evaluate the mechanisms underlying declines in mobility with aging among community-dwelling older adults. Our primary objective was to design and implement a CPET protocol in older adults that was dependable, safe, scientifically valuable, and methodologically reliable.

**METHODS:** CPET was performed using treadmill exercise in 875 adults ≥70 years. A composite protocol included a symptom-limited peak exercise phase and two submaximal phases to assess cardiopulmonary ventilatory indices during 1) participants’ preferred walking speed and 2) at slow walking speed of 1.5 mph (0.67 m/s). An adjudication process was in place to review tests for validity if they met any prespecified criteria (VO2peak <12.0 ml/kg/min; maximum heart rate (HR) <100 bpm; respiratory exchange ratio (RER) <1.05 and a rating of perceived exertion <15). A repeat test was performed in a subset (N=30) to assess reproducibility.

**RESULTS:** CPET was safe and well tolerated, with 95.8% of participants able to complete the VO2peak phase of the protocol. Only 56 (6.4%) participants had a risk alert during any phase of testing and only two adverse events occurred during the peak phase: a fall and atrial fibrillation. The average ± standard deviation for VO2peak was 20.2 ± 4.8 mL/kg/min, peak HR 142 ± 18 bpm, and peak RER 1.14 ± 0.09. VO2peak and RER were slightly higher in men than women. Adjudication was indicated in 47 participants; 20 were evaluated as valid, 27 as invalid (18 had a data collection error, 9 did not reach VO2peak). Reproducibility of VO2peak was high (intraclass correlation coefficient=0.97).

**CONCLUSIONS:** CPET was feasible, effective and safe for community-dwelling older adults, many of whom had multimorbidity and frailty. These data support a broader implementation of CPET to provide important insight into the role of CRF and its underlying determinants in aging and age-related conditions and diseases.

**Clinical Perspective:** *What Is New?:*

Performing cardiopulmonary exercise testing in a community dwelling older adult with multimorbidities or frailty is feasible and exceptionally safe under highly trained exercise physiologists and physician supervision.
Reproducibility of VO2peak among community-dwelling older adults with significant clinical complexity was high (intraclass correlation coefficient=0.97).
The VO2peak observed was comparable to established normative data for older adults, and adds merit to the limited data collected on VO2peak norms in older adults.

*What Are the Clinical Implications?:*

Ventilatory gas collection during clinical cardiac stress testing may be valuable to plan of care in routine management of older adults due to the important role of aerobic fitness on morbidity and mortality.
Cardiopulmonary exercise testing can provide insight into the role of cardiorespiratory fitness and its underlying determinants in aging and age-related conditions and diseases.

## INTRODUCTION

Cardiorespiratory fitness (CRF) refers to the capacity of the circulatory and respiratory systems to supply oxygen to skeletal muscle mitochondria for energy production needed during physical activity.^1^ Peak oxygen consumption (VO2peak) is the criterion measure of CRF, which declines with aging, is a key metric of physical function and a strong predictor of risk for morbidity and mortality.^2^ It is often applied to diagnose and evaluate the effects of disease as well as constitutive physiologic mechanisms.^3^ VO2peak assessed using symptom-limited cardiopulmonary exercise testing (CPET) is a vetted criterion metric of CRF.^1,4,5^ CPET entails an exercise provocation to stimulate integrated physiologic responses (cardiac, pulmonary, muscle, autonomic, circulatory) that can be assessed through patterns in ventilatory indices.^1,6^ Measurements of oxygen consumption (VO2), carbon dioxide production (VCO2), minute ventilation, and associated indices during progressive exercise intensity assess aerobic capacity, ventilatory efficiency, and other aspects of physical function. These can discern patterns of performance and physical limitation, with a wide range of clinical and research applications. The reliability of CPET for measuring CRF in younger populations has been well-established.^1,7^ Its application, feasibility and reliability in older adults, however, is less certain. Studies to validate VO2peak in older adults have been limited to small sample sizes, with many lingering questions regarding their feasibility, reliability and safety amidst age-related issues of mobility impairments, multimorbidity, frailty, fear of falling or maximal exertion.^8,9^

The conceptual value of CRF assessments using CPET in older adults is strong. VO2peak correlates with physical function^10–12^, frailty, disability, and mortality^13^ in older adults. CPET has been useful in determining the etiology of dyspnea, a common problem for older adults.^1^ Likewise, CPET is useful in determining diagnosis, prognosis and management of heart failure, chronic obstructive lung disease and other clinical challenges pertinent to older adults.^1^

In the Study of Muscle, Mobility and Aging (SOMMA), a study designed to distinguish subcellular mechanisms in skeletal muscle underlying mobility disability and functional decline over a three year period in older adults, CPET is integral to the primary study objectives to clarify key relationships between skeletal muscle mitochondrial respiration and CRF. Optimal accuracy of CPET is paramount to better understand these relationships.^14^ SOMMA and other studies of aging and age-related conditions and diseases require a CPET protocol that is dependable, safe and reliable.

Many rudimentary issues for exercise testing become more complex for older adults, especially those who have no prior experience with exercise testing or training. Goals to achieve adequate motivation and confidence for a high exercise workload are often difficult, especially as many older adults become fearful at higher exercise intensities. While CPET using a bicycle has advantages of stability and safety, cycling is not an exercise that is familiar to many older Americans. In comparison, walking is a more familiar exercise that may provide a more meaningful evaluation of physiologic capacities, but concerns regarding walking limitations (gait instability, pain) and safety (falls, anxiety, hemodynamics, arrhythmia) may also increase with treadmill modes.^15–17^ Similarly, while breathing through a mouthpiece is commonly employed for CPET, the discomfort associated with mouthpieces may become disproportionate due to changes in jaw strength and dentition, and difficulty tolerating a nose clip and breathing exclusively through the mouth. For many older adults, face masks provide greater comfort and feasibility during exercise. However, it is not clear if face masks can achieve effective air seals for accurate ventilatory measurements for those experiencing age-related changes in facial structure.^18^

The primary aim of this report was to provide a detailed overview of the SOMMA CPET protocol. We report methods to achieve dependable assessments of cardiorespiratory fitness (VO2peak) and submaximal exercise performance with associated safety and reliability. We also examine feasibility of CPET among subpopulations in SOMMA with functional limitations and comorbidities.

## METHODS

### SOMMA Participants

Details of the design of SOMMA are published elsewhere (https://sommaonline.ucsf.edu).^19^ Volunteers were recruited at two clinical sites (University of Pittsburgh and Wake Forest University School of Medicine) from April 2019 to December 2021.^19^ Individuals were eligible to participate if they were ≥70 years old, willing and able to complete a skeletal muscle biopsy and undergo magnetic resonance (MR). Individuals were excluded if they reported an inability to walk ¼ mile or climb a flight of stairs; had body mass index (BMI) ≥40 kg/m^2^; had an active malignancy or dementia; had medical contraindication to biopsy or MR. Finally, participants must have been able to complete the 400-meter walk; those who seemed unable to do so at the screening visit completed a 4-meter walk to ensure their walking speed was ≥0.6 m/s. All participants provided written informed consent, and the study was approved by the WIRB-Copernicus Group. The goal was to collect CPET data on all participants at the baseline visit. The SOMMA baseline data was collected over three separate visits, with a goal of completion within 30 days. The first visit included a 400-meter walk test, strength and physical function measures, a detailed medical review of medical history and a resting electrocardiogram (ECG). CPET was done at the second visit. This provided the CPET team several days to obtain any pertinent details of prior ECGs or medical history to ensure each participant’s safety to complete the anticipated CPET. Completion of the 400-meter walking test provided SOMMA participants with an initial experience of a monitored functional assessment that contributed to greater confidence for the later CPET.

### Rationale for CPET Protocol Selection

The CPET protocol was developed by a working group of exercise physiologists and investigators with expertise in studies of older adults. CPET in SOMMA was performed using treadmill exercise with the rationale that walking is a more universally familiar exercise to older adults than bicycling. Walking was also better aligned with functional capacities needed for independent mobility. The SOMMA CPET protocol was structured with 3 phases to capture a full range of functional domains: (Phase 1) preferred walking speed (PWS); (Phase 2) progressive exercise with increasing incline to evaluate VO2peak (Peak); (Phase 3) slow walking speed (SWS). The reasoning for a three-phase protocol was to observe peak aerobic capacity and incorporate other novel submaximal indices of walking energetics.^10^

CPET was conducted by experienced exercise physiologists under the supervision of a physician or physician assistant. Staff conducting CPET went through formal, centralized training with steps to standardize the testing process, including participant encouragement. Contraindications for exercise testing were consistent with standard American College of Sports Medicine (ACSM) guidelines.^20^ To further optimize safety, eligibility screening included an “alert system”, where potential safety concerns were systematically reviewed. These potential concerns, grouped as low and high risk, included identification of alerts from blood pressure (BP), heart rate (HR), and ECG that warranted further evaluation before CPET testing or, if occurring during CPET, stopping testing and evaluating clinically (Listed in Table 1). Pre-CPET low-risk alerts required clearance by the study physician for CPET to proceed or were not cleared for Phase 2, while high-risk alerts led to a hold until participants were cleared by their personal physician, the study physician and the study medical safety officer. Alerts during CPET were consistent with the ACSM guideline’s relative and absolute contraindications for stopping the test.^20^

**Table 1.**
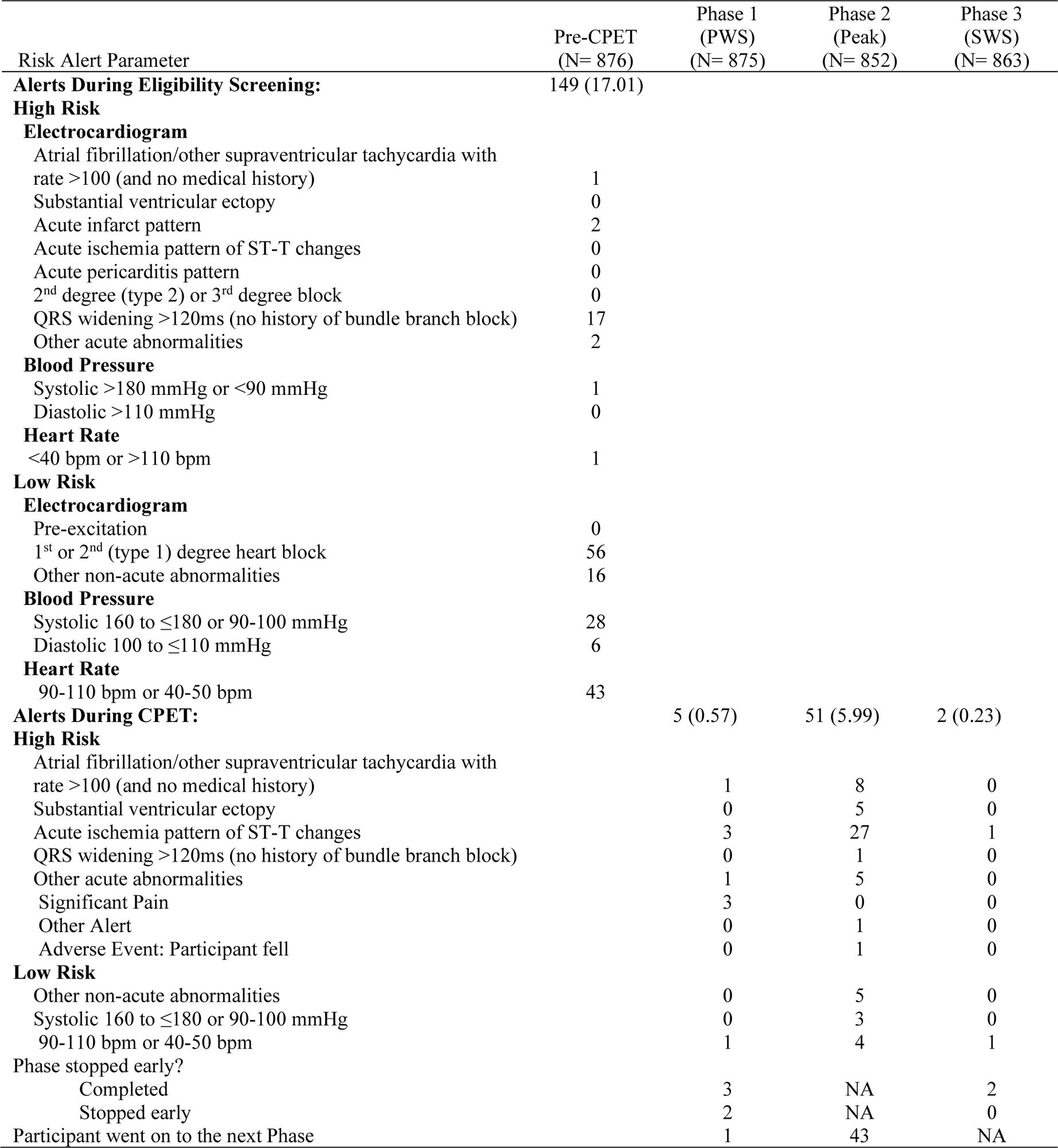
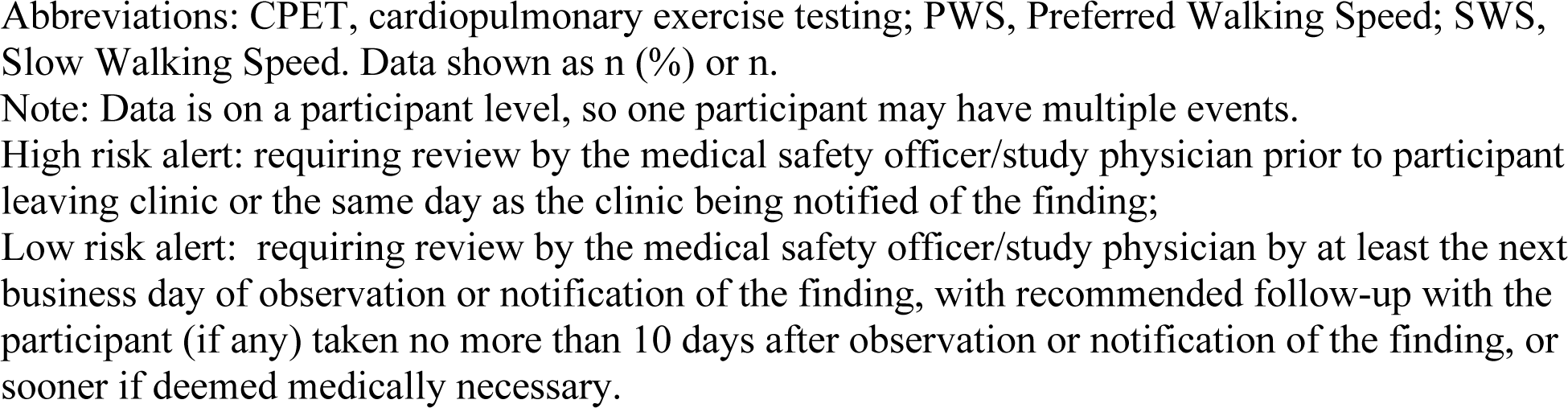
Risk Alerts occurring during CPET Eligibility or Testing: The Study of Muscle, Mobility and Aging (SOMMA)

| Risk Alert Parameter | Pre-CPET<br>(N= 876) | Phase 1<br>(PWS)<br>(N= 875) | Phase 2<br>(Peak)<br>(N= 852) | Phase 3<br>(SWS)<br>(N= 863) |
| --- | --- | --- | --- | --- |
| **Alerts During Eligibility Screening:** | 149 (17.01) |  |  |  |
| **High Risk** |  |  |  |  |
| **Electrocardiogram** |  |  |  |  |
| Atrial fibrillation/other supraventricular tachycardia with rate >100 (and no medical history) | 1 |  |  |  |
| Substantial ventricular ectopy | 0 |  |  |  |
| Acute infarct pattern | 2 |  |  |  |
| Acute ischemia pattern of ST-T changes | 0 |  |  |  |
| Acute pericarditis pattern | 0 |  |  |  |
| 2<sup>nd</sup> degree (type 2) or 3<sup>rd</sup> degree block | 0 |  |  |  |
| QRS widening >120ms (no history of bundle branch block) | 17 |  |  |  |
| Other acute abnormalities | 2 |  |  |  |
| **Blood Pressure** |  |  |  |  |
| Systolic >180 mmHg or <90 mmHg | 1 |  |  |  |
| Diastolic >110 mmHg | 0 |  |  |  |
| **Heart Rate** |  |  |  |  |
| <40 bpm or >110 bpm | 1 |  |  |  |
| **Low Risk** |  |  |  |  |
| **Electrocardiogram** |  |  |  |  |
| Pre-excitation | 0 |  |  |  |
| 1<sup>st</sup> or 2<sup>nd</sup> (type 1) degree heart block | 56 |  |  |  |
| Other non-acute abnormalities | 16 |  |  |  |
| **Blood Pressure** |  |  |  |  |
| Systolic 160 to ≤180 or 90-100 mmHg | 28 |  |  |  |
| Diastolic 100 to ≤110 mmHg | 6 |  |  |  |
| **Heart Rate** |  |  |  |  |
| 90-110 bpm or 40-50 bpm | 43 |  |  |  |
| **Alerts During CPET:** |  | 5 (0.57) | 51 (5.99) | 2 (0.23) |
| **High Risk** |  |  |  |  |
| Atrial fibrillation/other supraventricular tachycardia with rate >100 (and no medical history) |  | 1 | 8 | 0 |
| Substantial ventricular ectopy |  | 0 | 5 | 0 |
| Acute ischemia pattern of ST-T changes |  | 3 | 27 | 1 |
| QRS widening >120ms (no history of bundle branch block) |  | 0 | 1 | 0 |
| Other acute abnormalities |  | 1 | 5 | 0 |
| Significant Pain |  | 3 | 0 | 0 |
| Other Alert |  | 0 | 1 | 0 |
| Adverse Event: Participant fell |  | 0 | 1 | 0 |
| **Low Risk** |  |  |  |  |
| Other non-acute abnormalities |  | 0 | 5 | 0 |
| Systolic 160 to ≤180 or 90-100 mmHg |  | 0 | 3 | 0 |
| 90-110 bpm or 40-50 bpm |  | 1 | 4 | 1 |
| Phase stopped early? |  |  |  |  |
| Completed |  | 3 | NA | 2 |
| Stopped early |  | 2 | NA | 0 |
| Participant went on to the next Phase |  | 1 | 43 | NA |

Abbreviations: CPET, cardiopulmonary exercise testing; PWS, Preferred Walking Speed; SWS, Slow Walking Speed. Data shown as n (%) or n. Note: Data is on a participant level, so one participant may have multiple events. High risk alert: requiring review by the medical safety officer/study physician prior to participant leaving clinic or the same day as the clinic being notified of the finding; Low risk alert: requiring review by the medical safety officer/study physician by at least the next business day of observation or notification of the finding, with recommended follow-up with the participant (if any) taken no more than 10 days after observation or notification of the finding, or sooner if deemed medically necessary.

### CPET Equipment

Exercise was performed on a treadmill (Trackmaster, Full Vision Inc. Newton, KS) coupled with a cardiopulmonary metabolic cart (Medgraphics Ultima Series, Medgraphics Corporation, St. Paul, MN). Breath-by-breath ventilatory measurements were assessed using face masks (Hans Rudolph Inc., Shawnee, KS) fitted to each participant according to size options. Initially, an adjustable neoprene face mask that was designed to stretch over participants’ faces was used (Medgraphics, n=94). After discovering inconsistencies in ventilatory measurements from the neoprene mask, its use was discontinued and the Hans Rudolph mask was used. If a participant preferred, a mouthpiece and nose clip were substituted for the face mask (n=3 participants).

12-lead (Mason-Likar) ECG waveforms and pulse oxygen saturation (Nonin Inc., Plymouth, MN) were monitored continuously. HR, BP and rating of perceived exertion (RPE)^21^ were assessed in each of three phases of the CPET protocol.

### Familiarization

A familiarization period was performed prior to starting the CPET protocol. The goal of familiarization was to first demonstrate and then ensure the participant could achieve proper walking gait, and then explain each phase of the protocol. Exercise physiologists demonstrated these steps before encouraging the participants to repeat them and confirm successful learning. Familiarization included emphasis on the importance of normal stride length, safe positioning, regular breathing, and focusing eyes ahead. The speeds utilized at each phase were demonstrated, differentiating between PWS and SWS. Following the familiarization period, the participant was prepared with electrode lead placement and face mask fitting. Participants then sat for resting hemodynamics.

### The CPET Protocol

The CPET protocol was structured with three phases: (Phase 1) preferred walking speed (PWS); (Phase 2) progressive exercise with increasing incline to evaluate VO2peak (Peak); (Phase 3) slow walking speed (SWS). Phases 1 and 2 were conducted sequentially without interruption. After Phase 2, participants were given a 20-minute rest before beginning Phase 3.

Phase 1 consisted of 5 minutes of treadmill walking at each participant’s PWS with 0% incline. The PWS was based on the participant’s gait speed during the 400-meter walk test that was completed at the first SOMMA study visit. Continuous collection of ventilatory gases was performed as participants walked, along with assessments of RPE and manual BP at minutes 4 and 5.

The exercise physiologists used consistent coaching phrases to describe Phase 1 as a “warm-up” before the progressive intensity exercise of Phase 2 began. However, if a participant reached a peak effort during Phase 1 (n=4), exercise intensity would not increase, and CPET would progress directly to recovery part of Phase 2.

Phase 2 entailed a progression of exercise intensity to achieve VO2peak. The goals were to achieve a respiratory exchange ratio (RER) ≥1.05 and/or RPE ≥17. The VO2peak is defined as the highest 30-second average of VO2 achieved during Phase 2. The Peak protocol was premised on a Modified Balke protocol^17^ with systematized modifications to better suit the SOMMA population.

Whereas the Modified Balke protocol usually entails walking at either 2.0 or 3.0 mph (0.89 or 1.34 m/s), in SOMMA the initial treadmill speed was the PWS. If a participant’s PWS was <2.0 mph (0.89 m/s), the initial speed used in Phase 2 was increased, but with discretion by the exercise physiologist to select a speed that was well tolerated; the initial speeds used in Phase 2 were <2.0 mph (0.89 m/s) for 49 participants. Furthermore, the Modified Balke protocol usually starts at a 2.5% incline and increases 2.5% at each 2-minute stage thereafter until a participant reaches volitional fatigue. In SOMMA, 2.5% increases in treadmill incline were made every 2 minutes, but limited to a maximal incline of 10% to avoid biomechanically awkward walking at steeper angles. Thereafter, higher exercise intensity beyond the 10% incline relied on increasing treadmill speed up to 0.5 mph (0.22 m/s) at each 2-minute stage until volitional fatigue. To evoke a maximal effort, participants were prompted before each change in speed and incline and encouraged to continue until they could no longer maintain the given workload. Every 2 minutes and at peak performance, HR, BP and RPE were assessed.

Once the participant reached volitional fatigue, a 1-minute recovery period walking at 1 mph (0.45 m/s) and 0% incline was initiated, followed by 4 minutes of seated recovery. During the recovery periods continuous assessments of ventilatory indices, ECG, HR, and oxygen saturation were completed as well as monitoring of BP and RPE at minute 5. The participant was asked to continue sitting for the remainder of the 20-minute recovery period.

During Phase 3, the participant walked on the treadmill at 1.5 mph (0.67 m/s) and 0% incline, with continuous collection of ventilatory gases and assessments of RPE at minutes 4 and 5 and manual BP at minute 5.

Throughout each of the phases of testing, risk alerts associated with the exercise provocation were carefully monitored and addressed by the supervising exercise physiologist and clinician as per ACSM guidelines.^20^

### Quality Control

The CPET metabolic cart received maintenance every 6 months to ensure proper calibration (ventilatory, treadmill speed, grade). The CPET working group optimized the protocol and face mask used for testing, created a format for saving and sharing data, and created an adjudication process to review validity of tests in question. The CPET working group met regularly to review protocol questions, safety and data quality. Data collected were reviewed monthly by the working group to evaluate consistency between sites in the application of the protocol.

### Adjudication of VO2peak Data

To ensure that Phase 2 data were accurate, tests were adjudicated if they met at least one of the following criteria: VO2peak <12 ml/kg/min; maximum RPE <15 and maximum RER<1.05 (up to the start of recovery); maximum HR <100 bpm. Adjudicators were assigned from the CPET working group and were provided with data from CPET along with participant characteristics (age, sex, anthropometrics, gait speed, medication use). Adjudicators determined the validity of the Phase 2 data, as well as indicated if the data had a systematic problem and should not be used for the other two phases.

### Repeatability

Repeatability testing was conducted on a convenience sample of n=30 (15 per site, equal number by sex), who repeated all phases of the CPET protocol. The goal was for the repeat tests to be completed 7 days after the initial CPET, at the same time of day.

### Assessment of Walking Energetics

To examine walking energetics for both Phases 1 and 3 (PWS, SWS), the energetic cost of walking (ECW) was calculated as the average VO2 consumption from the last 3 minutes of each test. We derived the energy cost-capacity ratio of both phases as 100*ECW/VO2peak.^11^

### Participant Phenotypes and Characteristic Data

SOMMA recruited community-dwelling older adults with diverse characteristics. Multimorbidity, frailty and recurrent falling were quantified as it is often assumed that these subpopulations are unable to complete maximal exercise testing. Depression was defined as a score ≥10 on the Center for Epidemiological Studies Depression Scale (CES-D-10).^22^ Information on self-reported history of chronic health conditions and depression was combined to create the SOMMA multimorbidity index (0-11).^23^ The frailty phenotype was defined following the criteria by Fried et al.^24^ Participants reported information about lifestyle, falls history, mobility limitations, smoking status and medication use.^25,26^ Participants were categorized as recurrent fallers if they reported ≥2 falls in the past year. Self-reported physical activity was assessed with the Community Healthy Activities Model Program for Seniors (CHAMPS) questionnaire.^27^ Age predicted maximal heart rate (APMHR) was estimated as 208 -0.7 *age.^28^ Slow gait speed was defined as ≤0.8 m/s on the 4-meter test. Obesity was defined as BMI≥30 kg/m^2^.

### Statistical Analysis

Participant characteristics, phenotypic data and CPET phase completion were summarized by means and standard deviations (SD) for continuous variables, counts and percentages for categorical variables. Differences in these characteristics between participants cleared or excluded from the Phase 2 portion of CPET were analyzed using t-tests or Wilcoxon rank-sum tests for continuous variables, χ2 tests for categorical characteristics. Similar comparisons were performed by phenotype and mask type. Similarly, differences in CPET measures by sex was analyzed. The distribution of VO2peak across age was displayed graphically.

For the repeatability analysis, differences between CPET parameters from the original and repeat measures were examined using paired t-tests. Agreement between the two measures was examined with correlations and intraclass correlation coefficients (ICC) and 95% confidence intervals (CI), computed using a 2-way analysis of variance. A coefficient of variation (CV) was also calculated.^29^ Bland-Altman plots were presented to assess systematic bias in the differences in measurement of four parameters of Phase 2 (VO2peak, maximum RER, maximum HR, maximum RPE).^30^ Formal tests of systematic bias were performed using linear regression to examine whether the scatter in the Bland-Altman plots was heteroscedastic.^31^ A mixed models approach was used to examine the fixed effect of site and order of test (original vs. repeat).

Significance levels reported were 2-sided. Statistical analyses were performed using SAS software, version 9.4 (SAS Institute, Inc, Cary, NC).

## RESULTS

### Participant Characteristics

Of the 879 SOMMA participants, 875 performed CPET. Of the 4 who had no CPET performed, 2 were never screened for CPET, one was not cleared for testing, and one was cleared but was unable to use the treadmill. Of the 875 who attempted CPET, 84.8% were White; 59% were women; average age was 76.3 ± 5.0 years; and average BMI was 27.6 ± 4.6 kg/m^2^. The average 4-meter usual-pace gait speed was 1.04 ± 0.20 m/s. Only 41 (4.8%) had 3 or more chronic conditions. Based on CHAMPS, participants reported an average baseline of 7 hours per week of moderate-intensity physical activity (Table 2).

**Table 2.**
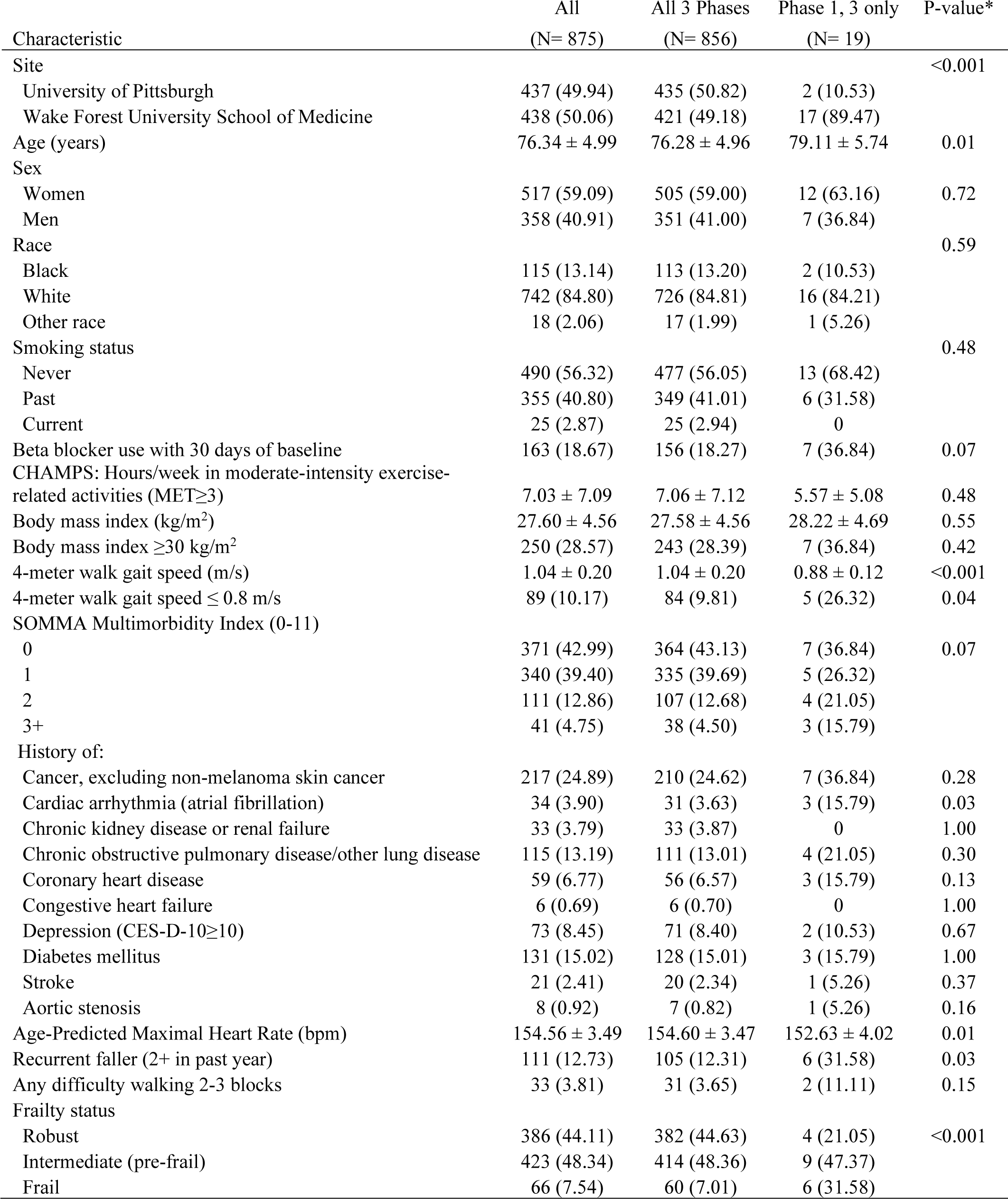
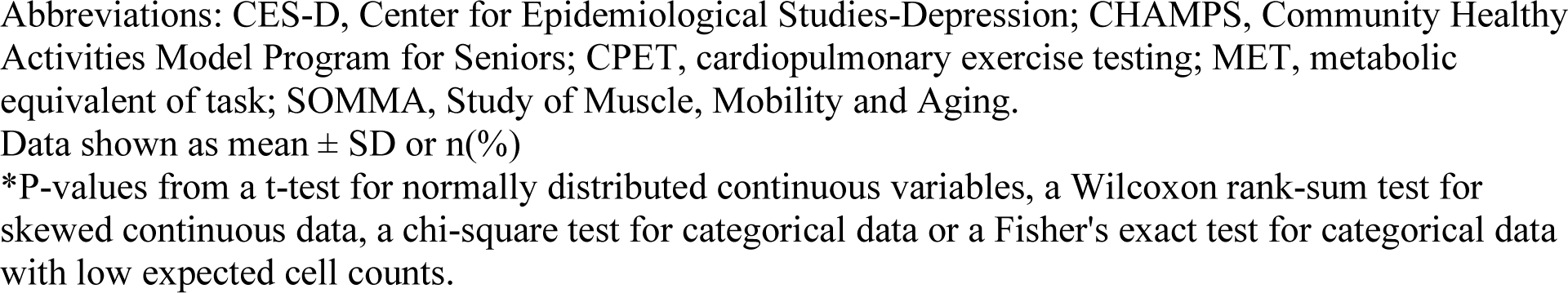
Baseline Characteristics by CPET Eligibility: The Study of Muscle, Mobility and Aging (SOMMA)

| Characteristic | All<br>(N= 875) | All 3 Phases<br>(N= 856) | Phase 1, 3 only<br>(N= 19) | P-value* |
| --- | --- | --- | --- | --- |
| Site |  |  |  | <0.001 |
| University of Pittsburgh | 437 (49.94) | 435 (50.82) | 2 (10.53) |  |
| Wake Forest University School of Medicine | 438 (50.06) | 421 (49.18) | 17 (89.47) |  |
| Age (years) | 76.34 ± 4.99 | 76.28 ± 4.96 | 79.11 ± 5.74 | 0.01 |
| Sex |  |  |  |  |
| Women | 517 (59.09) | 505 (59.00) | 12 (63.16) | 0.72 |
| Men | 358 (40.91) | 351 (41.00) | 7 (36.84) |  |
| Race |  |  |  | 0.59 |
| Black | 115 (13.14) | 113 (13.20) | 2 (10.53) |  |
| White | 742 (84.80) | 726 (84.81) | 16 (84.21) |  |
| Other race | 18 (2.06) | 17 (1.99) | 1 (5.26) |  |
| Smoking status |  |  |  | 0.48 |
| Never | 490 (56.32) | 477 (56.05) | 13 (68.42) |  |
| Past | 355 (40.80) | 349 (41.01) | 6 (31.58) |  |
| Current | 25 (2.87) | 25 (2.94) | 0 |  |
| Beta blocker use with 30 days of baseline | 163 (18.67) | 156 (18.27) | 7 (36.84) | 0.07 |
| CHAMPS: Hours/week in moderate-intensity exercise-related activities (MET≥3) | 7.03 ± 7.09 | 7.06 ± 7.12 | 5.57 ± 5.08 | 0.48 |
| Body mass index (kg/m<sup>2</sup>) | 27.60 ± 4.56 | 27.58 ± 4.56 | 28.22 ± 4.69 | 0.55 |
| Body mass index ≥30 kg/m<sup>2</sup> | 250 (28.57) | 243 (28.39) | 7 (36.84) | 0.42 |
| 4-meter walk gait speed (m/s) | 1.04 ± 0.20 | 1.04 ± 0.20 | 0.88 ± 0.12 | <0.001 |
| 4-meter walk gait speed ≤ 0.8 m/s | 89 (10.17) | 84 (9.81) | 5 (26.32) | 0.04 |
| SOMMA Multimorbidity Index (0-11) |  |  |  |  |
| 0 | 371 (42.99) | 364 (43.13) | 7 (36.84) | 0.07 |
| 1 | 340 (39.40) | 335 (39.69) | 5 (26.32) |  |
| 2 | 111 (12.86) | 107 (12.68) | 4 (21.05) |  |
| 3+ | 41 (4.75) | 38 (4.50) | 3 (15.79) |  |
| History of: |  |  |  |  |
| Cancer, excluding non-melanoma skin cancer | 217 (24.89) | 210 (24.62) | 7 (36.84) | 0.28 |
| Cardiac arrhythmia (atrial fibrillation) | 34 (3.90) | 31 (3.63) | 3 (15.79) | 0.03 |
| Chronic kidney disease or renal failure | 33 (3.79) | 33 (3.87) | 0 | 1.00 |
| Chronic obstructive pulmonary disease/other lung disease | 115 (13.19) | 111 (13.01) | 4 (21.05) | 0.30 |
| Coronary heart disease | 59 (6.77) | 56 (6.57) | 3 (15.79) | 0.13 |
| Congestive heart failure | 6 (0.69) | 6 (0.70) | 0 | 1.00 |
| Depression (CES-D-10≥10) | 73 (8.45) | 71 (8.40) | 2 (10.53) | 0.67 |
| Diabetes mellitus | 131 (15.02) | 128 (15.01) | 3 (15.79) | 1.00 |
| Stroke | 21 (2.41) | 20 (2.34) | 1 (5.26) | 0.37 |
| Aortic stenosis | 8 (0.92) | 7 (0.82) | 1 (5.26) | 0.16 |
| Age-Predicted Maximal Heart Rate (bpm) | 154.56 ± 3.49 | 154.60 ± 3.47 | 152.63 ± 4.02 | 0.01 |
| Recurrent faller (2+ in past year) | 111 (12.73) | 105 (12.31) | 6 (31.58) | 0.03 |
| Any difficulty walking 2-3 blocks | 33 (3.81) | 31 (3.65) | 2 (11.11) | 0.15 |
| Frailty status |  |  |  |  |
| Robust | 386 (44.11) | 382 (44.63) | 4 (21.05) | <0.001 |
| Intermediate (pre-frail) | 423 (48.34) | 414 (48.36) | 9 (47.37) |  |
| Frail | 66 (7.54) | 60 (7.01) | 6 (31.58) |  |

Abbreviations: CES-D, Center for Epidemiological Studies-Depression; CHAMPS, Community Healthy Activities Model Program for Seniors; CPET, cardiopulmonary exercise testing; MET, metabolic equivalent of task; SOMMA, Study of Muscle, Mobility and Aging. Data shown as mean ± SD or n(%) \*P-values from a t-test for normally distributed continuous variables, a Wilcoxon rank-sum test for skewed continuous data, a chi-square test for categorical data or a Fisher's exact test for categorical data with low expected cell counts.

Of the 875 cleared for CPET, 19 (2.2%) were only cleared for the two sub-peak phases (PWS, SWS). They were older, had slower average gait speed, more likely to have a history of atrial fibrillation, were more likely to be a recurrent faller and had a higher prevalence of frailty (p<0.05 for all, Table 2).

### Comparison of Face Mask Type

Comparison of data from participants by mask type showed that the Hans Rudolph masks achieved greater consistency of ventilatory measures than the neoprene face mask. VO2peak was similar for participants wearing either type of mask (p=0.29). However, participants wearing the Hans Rudolph mask were less likely to have their tests identified for adjudication (7.89% vs 17.02%, p<0.01). Of those participants with tests that needed adjudication, those wearing the Hans Rudolph mask were more likely to be adjudicated as valid (77.78% vs 25.00%, p<0.01). Few participants (n=6) had Phase 2 (Peak) stopped due to mask discomfort.

### Completion of the 3 Phases of CPET

As shown in Figure 1, there was a high rate of completion for the CPET protocol; 863 (98.6%) completed Phase 1 (PWS) and 860 (98.3%) completed Phase 3 (SWS). A total of 852 (97.3%) performed Phase 2 (Peak). Older participants were less likely to finish Phase 1 or Phase 2, but had similar completion rates to younger participants for Phase 3 (Supplemental Table 1). Compared to non-frail participants, those classified as frail were more likely to not be cleared for Phase 2 (9.09% vs 1.60%). Recurrent fallers were also more likely not to be cleared for Phase 2 (5.41% vs 1.70%). Recurrent fallers who completed the CPET protocol, however, had similar rates of risk alerts during the test as those who fell 0-1 times in the past year. Those “slow walkers” were more likely to be unable to complete Phase 2 (Supplemental Table 2). Those with 3 or more comorbidities were more likely to not have performed Phase 2 and were more likely to have stopped the protocol before reaching Phase 3. There were no differences in test completion by obesity (Supplemental Table 3).

**Figure 1.**
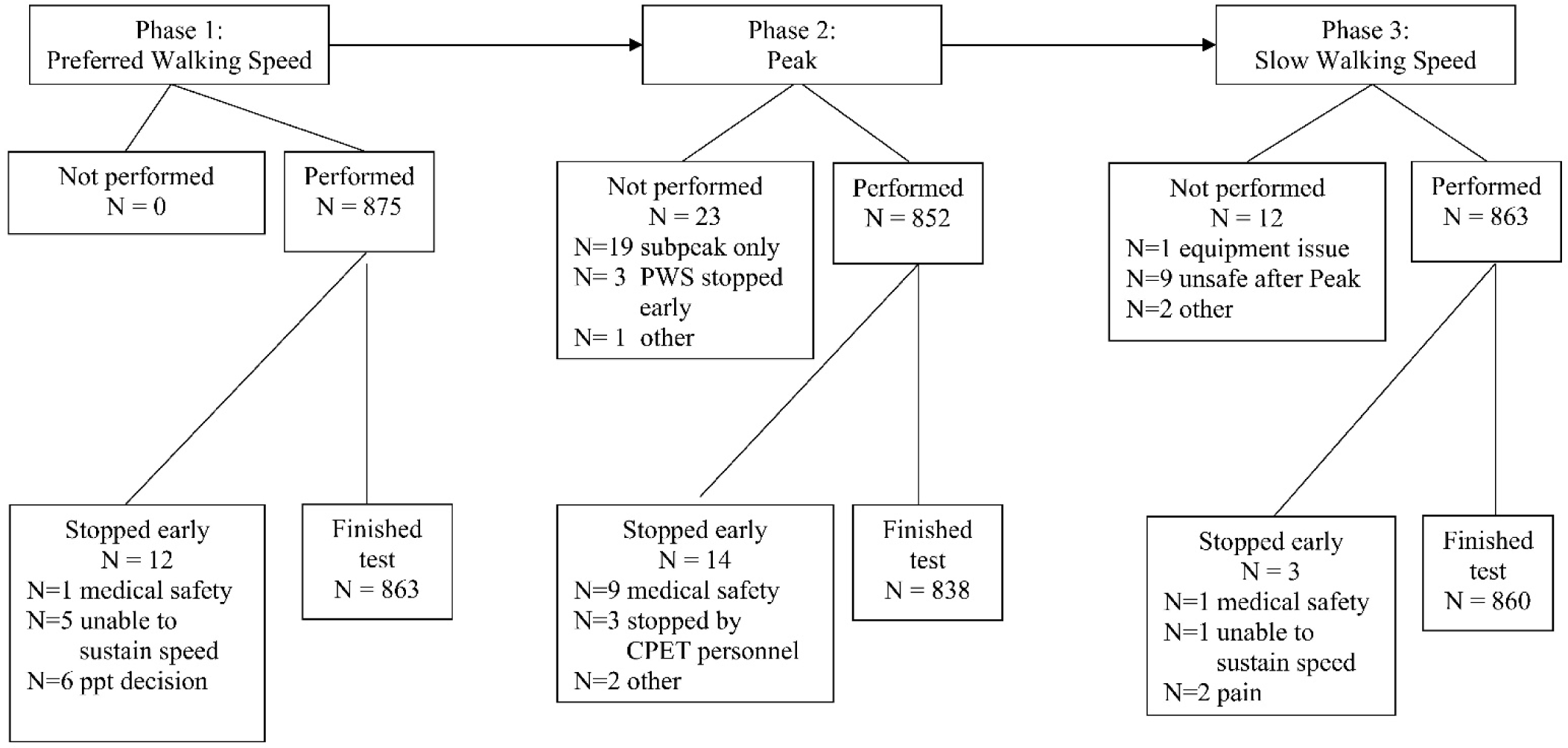
CPET performed in the Study of Muscle, Mobility and Aging (SOMMA)

### Pre-CPET Risk Alerts

Of the 876 screened for CPET eligibility, 189 (22%) had a low-risk alert during screening or CPET (Table 1). Blood pressure and ECG risk alerts during screening were common and were typically judged to be not serious such that the protocol was continued. Of the participants with a risk alert that occurred during screening, 94% were cleared for all 3 phases, and only 11% of those participants had an alert during CPET.

### Safety During CPET

Very few participants had a risk alert during Phases 1 and 3 [PWS 5 (0.6%), SWS 2 (0.2%)] (Table 1). Among 852 participants who attempted Phase 2 (Peak), 6% had a risk alert, although most went on to complete Phase 3. The most common alert during testing was acute ischemia pattern of ST-T changes (Phase 1, n=3; Phase 2, n=27; Phase 3, n=1). The medical safety officer was notified of all high risk alerts or adverse events occurring during CPET (n=50 participants). With the exception of 7 participants for whom the high risk alert was for a known condition, the remaining 43 participants were either contacted in person (by phone, email, letter), or were referred for medical treatment. There were two adverse events that occurred during Phase 2 (Peak). One participant fell on the treadmill. This adverse event resolved in one day with no need for treatment, and was classified as mild severity with minor skin irritation and soft tissue effects. One participant developed atrial fibrillation during the test and was referred to the emergency room for treatment, resulting in an overnight hospitalization. This event was considered serious, with resolution in one day. CPET for both these participants was stopped. There were no statistically significant differences in the number of risk alerts during CPET across age groups, frailty status, gait speed, multimorbidity status or obesity (Supplemental Tables 1-3).

### Ventilatory Indices

Men had higher VO2peak values on average than women (Table 3). The average maximum HR during Phase 2 (Peak) did not differ by sex. Men had a slightly higher maximum RER value during Phase 2 than women. (Figure 2). The average percentage of the maximum HR to the age-predicted maximum heart rate was 91.72 ± 11.42, with 60% reaching 90% or more of their age-predicted maximum heart rate.

**Figure 2:**
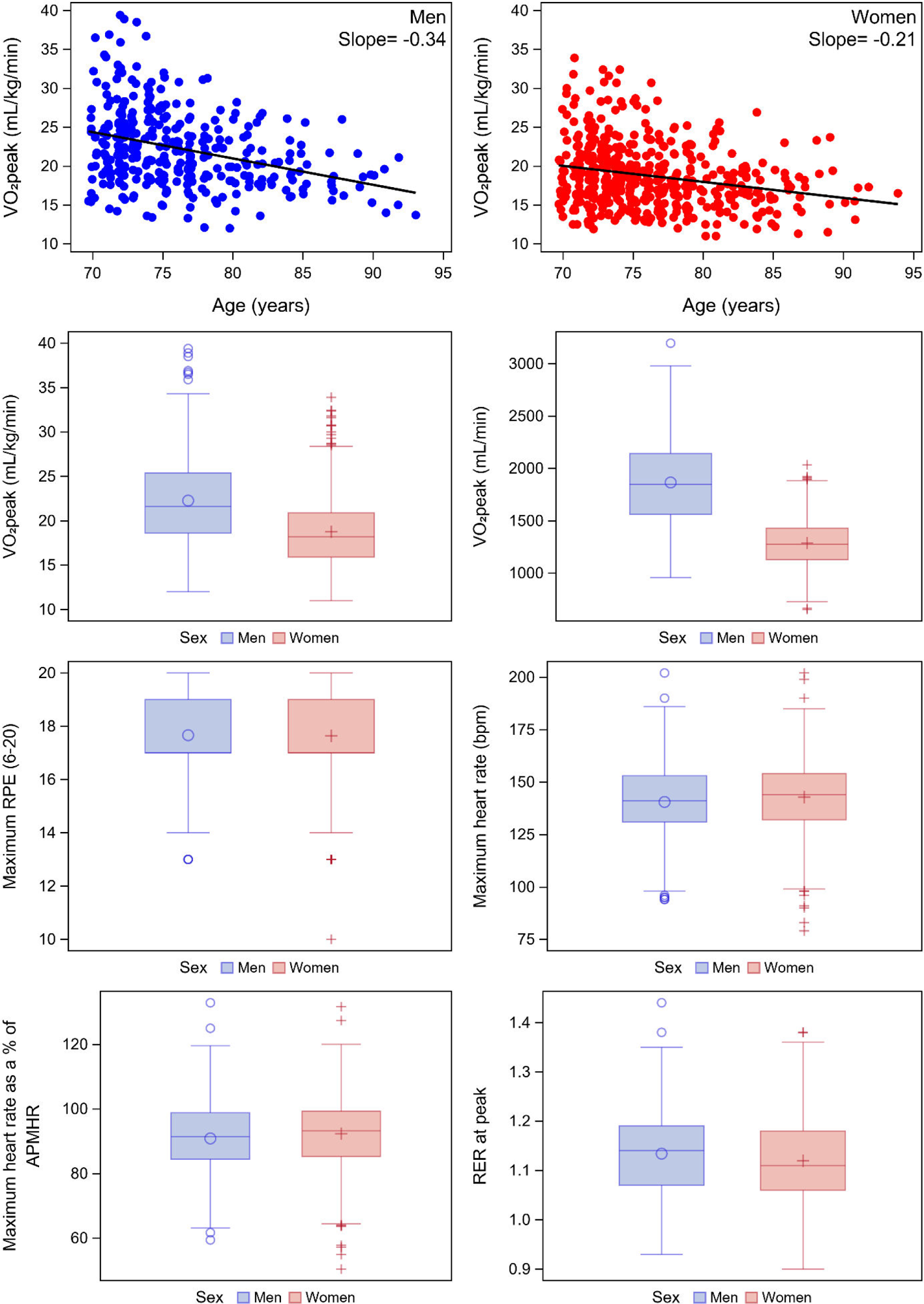
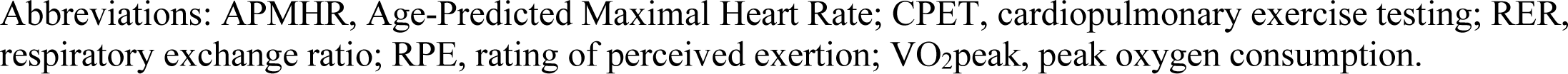
Distributions of CPET measures from Phase 2 (Peak) by Sex: The Study of Muscle, Mobility and Aging (SOMMA)

**Table 3.** CPET measures by sex: The Study of Muscle, Mobility and Aging (SOMMA)

Tables 3. CPET measures by sex: The Study of Muscle, Mobility and Aging (SOMMA)
| CPET Measure | All<br>(N= 875) | Men<br>(N= 358) | Women<br>(N= 517) | P-value* |
| --- | --- | --- | --- | --- |
| **Phase 1: Preferred Walking Speed** |  |  |  |  |
| Energetic cost of walking (mL/kg/min) | 12.44 ± 2.49 | 12.95 ± 2.47 | 12.07 ± 2.44 | <.0001 |
| Rating of Perceived Exertion (6-20) | 10.50 ± 2.03 | 10.32 ± 2.03 | 10.64 ± 2.02 | 0.02 |
| Heart rate (bpm) | 103.74 ± 17.22 | 99.35 ± 16.48 | 106.82 ± 17.08 | <.0001 |
| Respiratory exchange ratio | 0.90 ± 0.09 | 0.90 ± 0.09 | 0.89 ± 0.09 | 0.11 |
| Treadmill speed (m/s) | 1.05 ± 0.18 | 1.08 ± 0.17 | 1.02 ± 0.18 | <.0001 |
| Cost-Capacity Ratio (%) | 63.21 ± 12.39 | 59.90 ± 12.96 | 65.61 ± 11.39 | <.0001 |
| **Phase 2: Peak** |  |  |  |  |
| VO<sub>2</sub>peak (mL/kg/min) | 20.22 ± 4.82 | 22.27 ± 5.00 | 18.75 ± 4.11 | <.0001 |
| VO<sub>2</sub>peak (mL/min) | 1528.30 ± 433.40 | 1865.62 ± 410.69 | 1286.95 ± 248.94 | <.0001 |
| Maximum Rating of Perceived Exertion (6-20) | 17.65 ± 1.42 | 17.66 ± 1.37 | 17.64 ± 1.46 | 0.91 |
| Maximum heart rate (bpm) | 141.87 ± 18.00 | 140.50 ± 18.24 | 142.82 ± 17.79 | 0.07 |
| Maximum heart rate as a percentage of APMHR | 91.72 ± 11.42 | 90.85 ± 11.59 | 92.33 ± 11.27 | 0.06 |
| Respiratory exchange ratio at peak | 1.13 ± 0.09 | 1.13 ± 0.09 | 1.12 ± 0.09 | 0.02 |
| Maximum Respiratory exchange ratio | 1.14 ± 0.09 | 1.15 ± 0.08 | 1.14 ± 0.09 | 0.04 |
| **Phase 3: Slow Walking Speed (1.5 mph)** |  |  |  |  |
| Energetic cost of walking (mL/kg/min) | 9.46 ± 1.64 | 9.61 ± 1.59 | 9.35 ± 1.66 | 0.02 |
| Rating of Perceived Exertion (6-20) | 8.41 ± 1.90 | 8.18 ± 1.70 | 8.57 ± 2.01 | 0.008 |
| Heart rate (bpm) | 100.08 ± 15.78 | 96.73 ± 15.41 | 102.40 ± 15.63 | <.0001 |
| Respiratory exchange ratio | 0.80 ± 0.06 | 0.80 ± 0.06 | 0.80 ± 0.07 | 0.13 |
| Cost-Capacity Ratio (%) | 48.81 ± 12.56 | 44.90 ± 11.22 | 51.61 ± 12.73 | <.0001 |

Abbreviations: APMHR, Age-Predicted Maximal Heart Rate; CPET, cardiopulmonary exercise testing; PWS, Preferred Walking Speed; SWS, Slow Walking Speed; VO<sub>2</sub>, oxygen consumption; VO<sub>2</sub>peak, peak oxygen consumption.
Data shown as mean ± SD or n(%)
\*P-values from a t-test for normally distributed continuous variables, a Wilcoxon rank-sum test for skewed continuous data.

The average treadmill speed for Phase 1 (PWS) was 1.05 ± 0.18 m/s, with an average ECW of 12.44 ± 2.49 mL/kg/min, and a cost-capacity ratio of 63.21% ± 12.39%. On average, men had higher PWS and ECW, and lower cost-capacity ratio than women.

For Phase 3 (SWS) the average ECW was 9.46 ± 1.64 mL/kg/min, and an average cost-capacity ratio of 48.81 ± 12.56. On average, men had higher ECW and lower cost-capacity ratio than women.

For all but recurrent fallers, the average VO2peak was significantly lower among those in the compromised group (Supplemental Figure 1). Those 85 years and older had a lower average VO2peak by 2.87 mL/kg/min compared to younger participants, those classified as frail had a lower average VO2peak by 4.62 mL/kg/min compared to non-frail participants, those slow walkers had a lower average VO2peak by 4.49 mL/kg/min compared to faster walkers, those with ≥3 comorbidities had a lower average VO2peak by 3.23 mL/kg/min compared to those with 0-2 comorbidities, and obese participants had a lower average VO2peak by 3.50 mL/kg/min compared to those with BMI<30 kg/m^2^. VO2peak declined with age (Figure 2).

### Adjudication Process

Of the 852 participants that performed Phase 2 (Peak), 47 (5.5%) were flagged for adjudication. (Figure 3) The majority (n=28) were due to low VO2peak (VO2 <12 mL/kg/min), while 21 had low HR (maximum HR<100 bpm). Of the 47 adjudicated, data were deemed valid for 20 (42.6%). Nine (19.1%) were adjudicated as invalid because the participant was not able to complete the test. Eighteen (38.3%) were adjudicated as invalid, primarily due to technical or other unknown issues with the gas exchange data.

**Figure 3.**
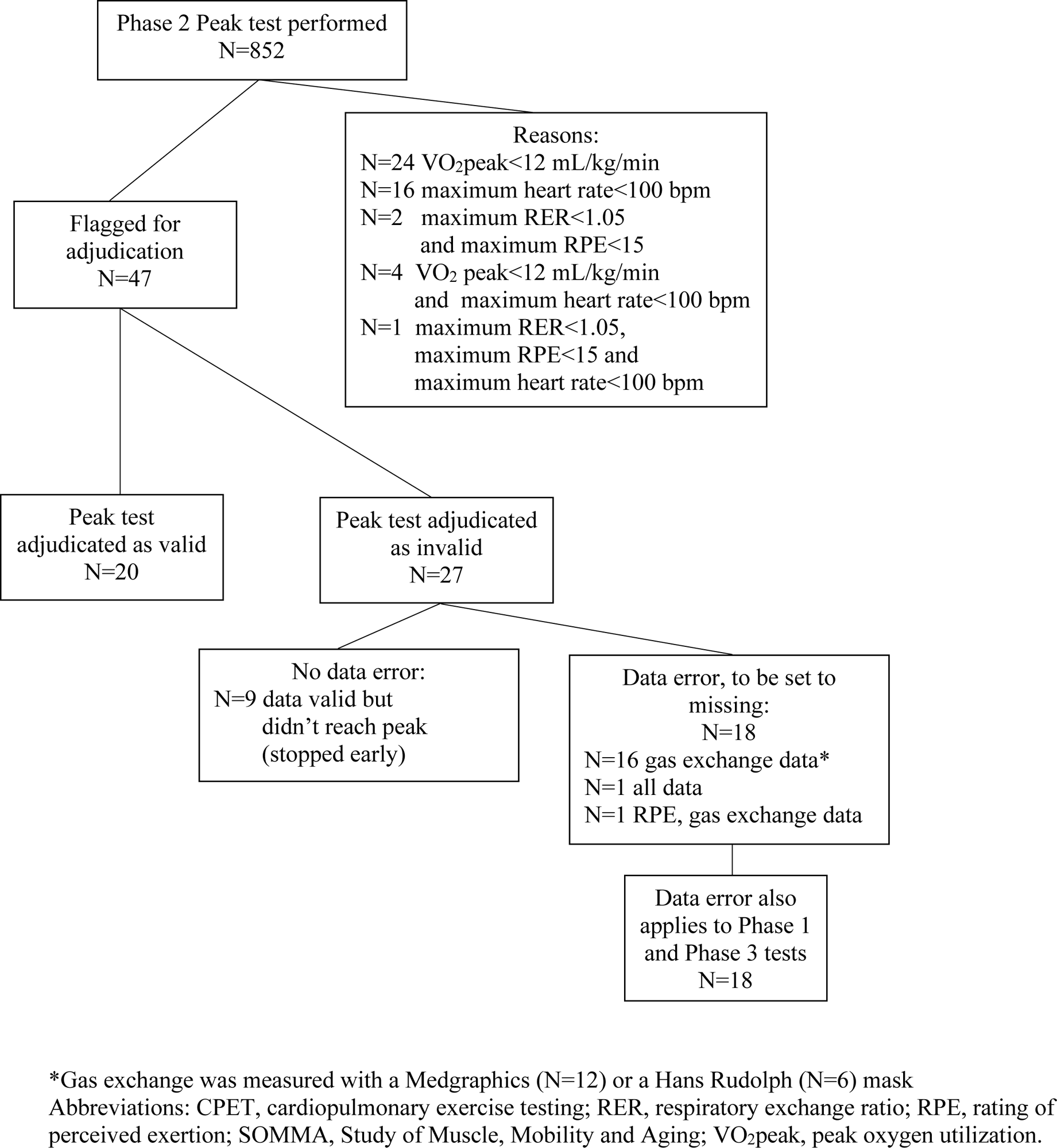
Adjudication Process for CPET data in the Study of Muscle, Mobility and Aging (SOMMA)

### Repeatability

The measurement of the Phase 2 (Peak) was highly reproducible (Table 4). The reliability as shown by the ICC demonstrated almost perfect agreement for VO2peak (0.97), and substantial agreement for maximum HR, RER and RPE (0.74 - 0.86). The Bland-Altman plots for these measures (Figure 4) illustrate a random scattering of points above and below the average mean difference with few points falling outside the 95% CI, showing a lack of systematic bias between the first and second measurements. The formal tests of heteroscedasticity were not significant (p>0.05). Results from mixed models for the Phase 2 measures did not show systematic bias based on study site (p >0.07) or whether the measurement was from the original or repeat measurement (p > 0.09).

**Figure 4.**
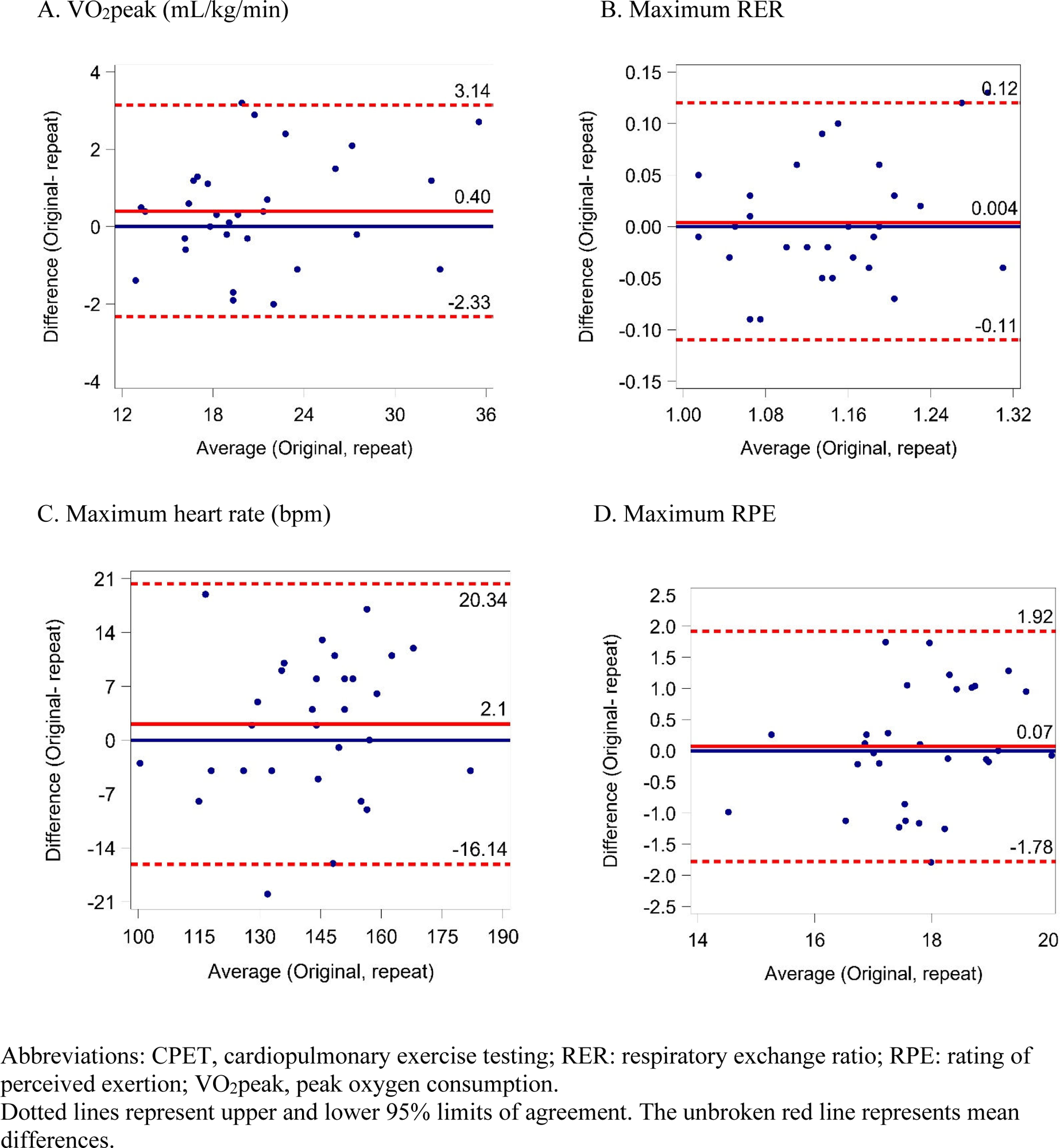
Bland-Altman plots showing the agreement between the original and repeat CPETs: The Study of Muscle, Mobility and Aging (SOMMA)

**Table 4.**
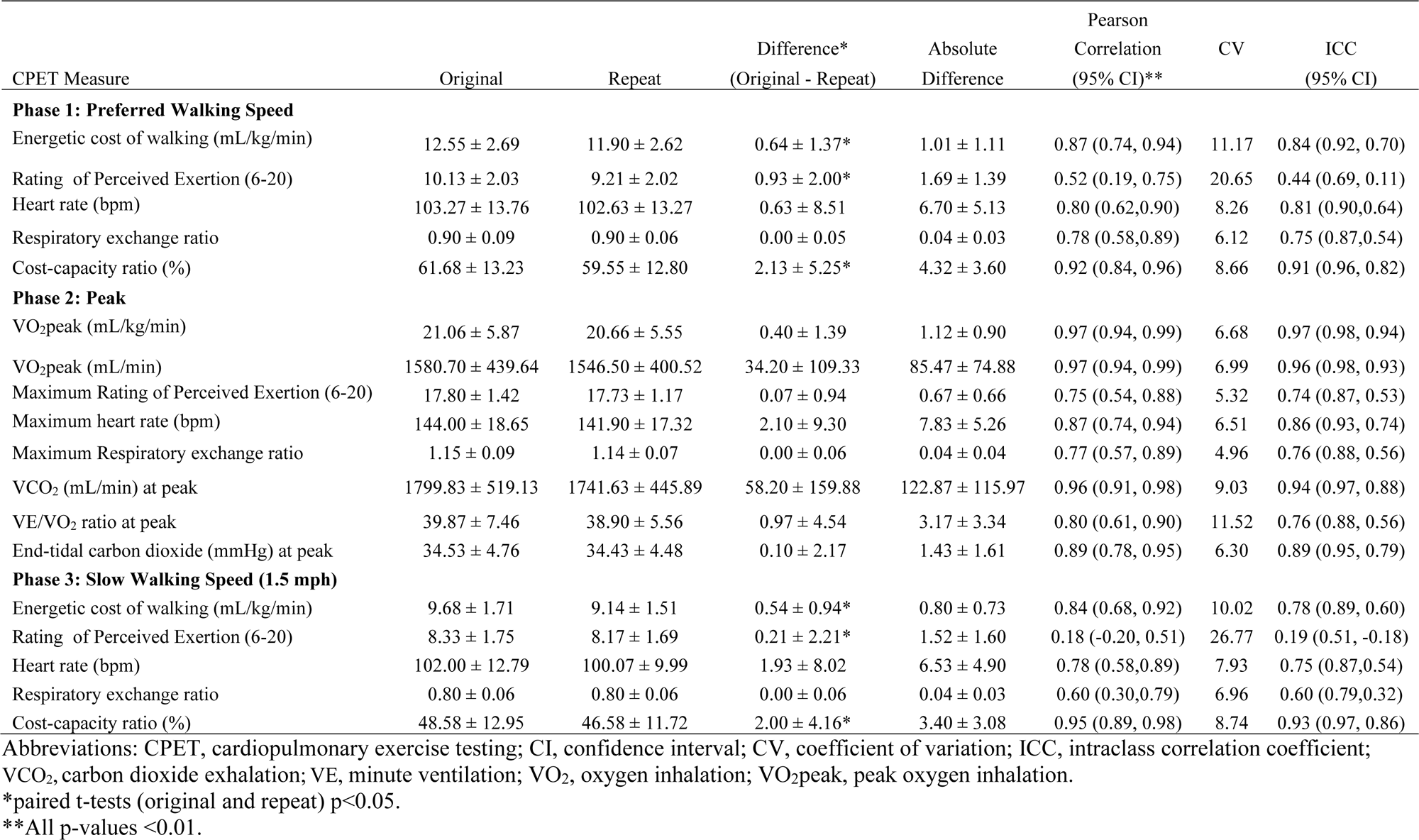
CPET repeatability (n=30): The Study of Muscle, Mobility and Aging (SOMMA)

Likewise, the reliability of ECW for Phases 1 and 3 (PWS, SWS) had strong agreement (ICCs 0.84, 0.78), but a poor agreement for RPE (ICCs 0.44, 0.19). Paired t-tests between the original and repeat measures from these phases showed a significant difference for ECW, RPE and cost-capacity ratio, with the original measurements being higher on average than the repeat measures. Results from mixed models did not show any systematic bias based on study site (p >0.09), but did show a systematic bias for whether the measurement was from the original or repeat measurement for ECW and cost-capacity ratio for both phases, and RPE for Phase 1 (p <0.05).

## DISCUSSION

The primary finding from this study is that CPET was highly feasible and safe to effectively obtain VO2peak as the criterion measure of CRF among older community-dwelling adults, including many with phenotypic features of frailty, multimorbidity, and poor physical function. Of the 879 participants enrolled in SOMMA, 99.7% were deemed eligible for CPET, and of those, 97.3% completed the peak exercise protocol. After adjudication, we obtained valid VO2peak values in 96.2% who attempted the test. Only 25 participants terminated CPET early. While others have reported results from CPET in older adults, they have been from smaller studies or conglomerations of multiple studies ^32–35^; and most lacked details regarding participant selection, or excluded participants with poor physical function, frailty or multimorbidity. Therefore, our study provides more generalizable data on the feasibility, safety and efficacy of CPET across a wider range of older adults.

The utility of CPET for older adults has often been questioned, largely because of safety concerns or reservations that many older adults cannot achieve peak exercise thresholds^15,36^ and/or that they are unable to achieve sufficient motivation or stability to achieve high exercise workloads. In SOMMA, we created a working group to design the CPET protocol specifically for older adults. We implemented a familiarization protocol, protocol standardization, quality control, safety alerts, and an adjudication process to assess valid VO2peak tests. Because of these rigorous methods, we obtained valid ventilatory measurements and had few risk alerts along with a very high proportion of individuals who reliably achieved VO2peak. This is particularly important for multicenter, longitudinal studies.

One of the key findings is that CPET is feasible and safe in community-dwelling older adults with a range of disease risk, disease burden and physical function when those with active cardiopulmonary disease and very slow gait speed have been excluded. Although procedures were implemented to exclude those in whom maximal exertion exercise was contraindicated, only 1 participant was not cleared for any phase of CPET and 19 (2%) were not cleared for Phase 2 (Peak), with a remarkably high proportion of participants having completed Phase 2 without incident. We monitored potential risk and uncovered a relatively small number of symptoms and ECG abnormalities. There were only two adverse events, one considered mild severity.

There were no significant differences in the number of risk alerts during CPET across age groups, frailty status, gait speed, multimorbidity status or obesity. Only 12.5% of alerts during CPET were from previously known conditions. Notably, 29% of participants with risk indicators during provocative exercise testing had a pre-CPET risk alert, suggesting high clinical value of CPET, as well as predominant safety of the assessment. Our results confirm the utility of CPET for ventilatory assessments in older adults, as well as for HR, BP, ECG, and other valuable nonventilatory physiology.^13^

These SOMMA CPET data also provide an opportunity to evaluate additional metrics, including submaximal respiratory data. The SOMMA cohort included participants at the lower end of physical function in which CPET data are limited. The safety of CPET has been reported previously, mostly in patients with cardiovascular and pulmonary disease^37^, but also among generally healthy adults.^35^ Still, less is known about the safety of CPET specifically in older adults, especially those with compromised physical function. Participant selection criteria, the diversity of populations studied and the CPET protocol all make it difficult to compare CPET safety across studies, yet our study strengthens rationale and method for application of CPET in older adults.

The average and range of VO2peak values in SOMMA are consistent with other databases and smaller studies in older adults.^33–35^ The recently published Fitness Registry and the Importance of Exercise: A National Data Base (FRIEND) showed VO2peak ranged from 13.6-29.4 ml/kg/min in septuagenarian men and 12.3-22.8 ml/kg/min in septuagenarian women.^35^ As in SOMMA, CRF is lower in older versus younger adults, and lower in women versus men, and there is heterogeneity among those being tested. SOMMA stands out in comparison to these reports by clarifying the functional impact of multimorbidity, frailty, obesity and other factors that are rarely assessed in CPET analyses. Future SOMMA analyses will explore in depth the associations of CPET with these factors. Reliable measures of VO2peak will be crucial to understand the mechanism underlying CRF and physical function. For example, we have reported in SOMMA that VO2peak is strongly associated with skeletal muscle mitochondrial energetics.^38^ Longitudinal SOMMA data will be instrumental in determining a role for CRF in mobility decline.

Repeated CPET showed excellent reproducibility for VO2peak. These data are consistent with previous reports which showed similarly strong test-retest reproducibility.^39,40^ The low RPE ICC during PWS and SWS may be attributed to a familiarization bias. There were no systematic site differences for any of the measures across all phases, showing consistency between multiple operators at multiple sites. Given that SOMMA entails treadmill exercise, the reproducibility data are even more notable.

### Limitations

Race and ethnic subgroups were not large enough to assess the differences of race/ethnicity. Further, SOMMA excluded candidates who could not complete the 400-meter walk or who walked <0.6 m/s over 4 meters, so results cannot be extended to those with more significant mobility disability.

### Conclusions

As a critical part of SOMMA we employed a CPET protocol that is feasible, safe, and effective in older adults with a wide range of physical function and disease, and could be used reliably as part of a multicenter study. SOMMA achieved high quality assessments of peak performance as well as submaximal assessments of walking efficiency. CPET data will provide a critical window into potential mechanisms underlying the biological aging process and risk for functional decline and morbidity. Moreover, implications of the SOMMA CPET protocol imply broader clinical and research potential, with generalizable efficacy for clinical care and research assessments among older adults. SOMMA uniquely demonstrates the value and utility of CPET, with potential to better understand and moderate organ and cellular changes that are associated with aging.

## ARTICLE INFORMATION

## Data Availability

Data is available at https://sommaonline.ucsf.edu

## Acknowledgements

CPET Working Group: Elvis A. Carnero; Steve Anthony; Cheyenne Barnett; Terri Blackwell; Peggy Cawthon; Amelia Cervantes; Robin Collins; Michelle Danielson; Daniel Forman; Nancy Glynn; Bret Goodpaster; Michelle Gordon; Teresa Harnish; Eileen Johnson; Justin Johnson; Kim Kennedy; Reagan Moffit; Anne Newman; Barbara Nicklas; Benjamin Schumacher; Frederico G. S. Toledo; April Tuttle; Katey Webber; Cody Wolf.

## Sources of Funding

The Study of Muscle, Mobility and Aging is supported by funding from the National Institute on Aging, grant number AG059416.” Study infrastructure support was funded in part by NIA Claude D. Pepper Older American Independence Centers at University of Pittsburgh (P30AG024827) and Wake Forest University (P30AG021332) and the Clinical and Translational Science Institutes, funded by the National Center for Advancing Translational Science, at Wake Forest University (UL1 0TR001420).

## Disclosures

All authors report no conflicts.

## Non-standard Abbreviations and Acronyms

ACSM: American College of Sports Medicine
APMHR: Age-Predicted Maximal Heart Rate BMI body mass index
BP: blood pressure
CES-D: Center for Epidemiological Studies-Depression
CHAMPS: Community Healthy Activities Model Program for Seniors
CI: confidence interval
CPET: cardiopulmonary exercise testing
CRF: cardiorespiratory fitness
CV: coefficient of variation
ECG: Electrocardiogram
ECW: energetic cost of walking
HR: heart rate
ICC: intraclass correlation coefficient
MET: metabolic equivalent of task
MR: magnetic resonance
PWS: preferred walking speed
RER: respiratory exchange ratio
RPE: rating of perceived exertion
SD: standard deviation
SOMMA: Study of Muscle, Mobility and Aging
SWS: slow walking speed
VCO2: carbon dioxide exhalation
VE: minute ventilation
VO2: oxygen inhalation
VO2peak: peak oxygen consumption

## Notes

### Competing Interest Statement

The authors have declared no competing interest.

### Clinical Trial

NA

### Author Declarations

All participants provided written informed consent, and the study was approved by the WIRB-Copernicus Group.

